# Multi-tissue analyses of allele-specific chromatin accessibility nominate likely functional variants for type 2 diabetes

**DOI:** 10.64898/2026.07.14.26358094

**Authors:** Narisu Narisu, Hannah X. Li, Caleb J. M. Rathbun, Arushi Varshney, Amy J. Swift, Tingfen Yan, Neelam Sinha, Kevin W. Currin, Dongxiang Xue, Catherine C. Robertson, D. Leland Taylor, Henry J. Taylor, Aimee Beck, Brian N. Lee, Li Wang, K. Alaine Broadaway, Emma P. Wilson, Heather Stringham, Jouko Saramies, Timo A. Lakka, Cassandra N. Spracklen, Laura J. Scott, Michael L. Stitzel, Jaakko Tuomilehto, Markku Laakso, Heikki A. Koistinen, Michael Boehnke, H. Efsun Arda, Shuibing Chen, Leslie G. Biesecker, Lori L. Bonnycastle, Michael R. Erdos, Karen L. Mohlke, Stephen C. J. Parker, Francis S. Collins

**Affiliations:** Center for Precision Health Research, National Human Genome Research Institute, National Institutes of Health, Bethesda, MD 20892, USA; Gilbert S. Omenn Department of Computational Medicine and Bioinformatics, University of Michigan, Ann Arbor, MI 48109, USA; Department of Genetics, University of North Carolina, Chapel Hill, NC 27599, USA; Department of Surgery, Center for Genomic Health, Weill Cornell Medicine, New York, NY 10065, USA; Department of Biology, Johns Hopkins University, Baltimore, MD 21218, USA; Department of Biostatistics and Center for Statistical Genetics, University of Michigan, Ann Arbor, MI, USA; South Karelia Social and Health Care District, Wellbeing Services County of South Karelia, Finland; Institute of Biomedicine, School of Medicine, University of Eastern Finland, Kuopio, Finland; Kuopio Research Institute of Exercise Medicine, Kuopio, Finland; Department of Clinical Physiology and Nuclear Medicine, Kuopio University Hospital, Kuopio, Finland; Department of Biostatistics and Epidemiology, University of Massachusetts Amherst, Amherst, MA 01003, USA; The Jackson Laboratory for Genomic Medicine, Farmington, CT, USA; Finnish Institute for Health and Welfare, Helsinki, Finland; Department of Public Health, University of Helsinki, Helsinki, Finland; Institute of Clinical Medicine, Internal Medicine, University of Eastern Finland, Kuopio, Finland; Faculty of Medicine, Research Programs Unit, Clinical and Molecular Metabolism (CAMM), University of Helsinki, Helsinki, Finland; Department of Medicine, University of Helsinki and Helsinki University Hospital, Helsinki, Finland; Minerva Foundation Institute for Medical Research, Helsinki, Finland; Laboratory of Receptor Biology and Gene Expression, Center for Cancer Research, National Cancer Institute, National Institutes of Health, Bethesda, MD 20892, USA

## Abstract

Genome-wide association studies (GWAS) have identified >1,200 signals associated with type 2 diabetes (T2D), yet identifying functional variants remains challenging because the majority of them lie in noncoding regions of the genome and are in areas of high linkage disequilibrium (LD). While chromatin accessibility QTL (caQTL) and expression QTL (eQTL) analyses are useful for nominating regulatory mechanisms underlying GWAS signals, limitations still exist in pinpointing functional variants within regions of high LD. A complementary approach that has been less frequently applied is to focus on the allele-specific effect on chromatin accessibility at heterozygous single-nucleotide polymorphisms (SNPs), hereafter referred to as “allelic imbalance”. We analyzed the allelic imbalance of reads generated from an assay for transposase-accessible chromatin with sequencing (ATAC-seq) across genotyped samples from 490 donors in T2D-relevant tissues: skeletal muscle, liver, pancreatic islets, adipose tissue, and relevant cell types. We identified 119,949 allelically imbalanced SNPs (FDR<0.05) across the genome. The allelic imbalance was often most prominent in one tissue and showed an enrichment overlapping with tissue-specific transcription factor (TF) binding footprints. Focusing on the 8,581 SNPs in previously published 99% credible sets from 338 T2D GWAS signals, we identified 256 imbalanced SNPs across 123 (36.4% of) signals, each showing allelic imbalance in at least one tissue or cell type. Of these, 71 signals contained only a single imbalanced SNP, representing excellent candidate causative variants. As a proof-of-concept, we showed that 23 of the 256 imbalanced SNPs were supported by allelic assays from previous studies. Further, we experimentally validated two imbalanced SNPs as likely functional variants: rs34584161 among a seven-SNP T2D credible set at the *RNF6* signal in islets and rs849134 among a 13-SNP credible set at the *JAZF1* signal in liver. This study demonstrates the power of integrating ATAC-seq allelic imbalance (ASAI) with GWAS statistical fine-mapping to identify candidate functional regulatory variants from among tightly linked GWAS variants in disease-relevant tissues. While applied here in T2D, this approach represents a widely applicable high-throughput framework for refining the genetic architecture of complex traits.

## Introduction

Type 2 diabetes (T2D) is a growing global health burden driven by both environmental and genetic factors^1–4^. Identifying causal genes and genetic variants is crucial for advancing our understanding of the molecular mechanisms underlying disease risk and progression, and helping to identify new strategies for disease stratification, prevention, and treatment.

Genome-wide association studies (GWAS) have identified >1,200 T2D association signals, encompassing thousands of single nucleotide polymorphisms (SNPs) and other types of genetic variants^1–4^. Linkage disequilibrium (LD) blocks in the human genome often span tens of kilobases; their existence made it possible to identify DNA regions with trait association with a limited genotyping panel in the early stages of GWAS^5,6^. However, LD poses a challenge in pinpointing the precise functional variants in signals with multiple tightly linked SNPs. For each association signal, statistical fine-mapping can be implemented to reduce the number of likely causal candidates to a “credible set.” Each SNP in the credible set is assigned a posterior probability of being causal^1–4^. Often credible sets are chosen to have an estimated 99% probability of including the causative SNP. Despite this effort, in the Mahajan *et al* study (2022) where such credible sets are published, 290 out of 338 of T2D GWAS credible sets (85.8%) still contain multiple SNPs in high LD^1^.

Demonstrating the effect of a causative SNP is often considered most robust when allelic differences are shown in an experimental assay. Existing approaches such as massively parallel reporter assays (MPRA), electrophoretic mobility shift assays (EMSA), luciferase gene reporter assays, and CRISPR editing have identified a modest number of functional T2D SNPs^7–12^, but these methods are typically costly, labor-intensive, and/or low-throughput. This highlights the need for efficient genome-wide methods to identify likely functional SNPs to prioritize for experimental validation.

The majority of fine-mapped GWAS loci lie in noncoding regions^13^, where candidate functional SNPs are thought to act through gene regulatory mechanisms. Chromatin accessibility quantitative trait locus (caQTL) analyses have identified genetic variants associated with differences in chromatin accessibility measured by assay for transposase-accessible chromatin with sequencing (ATAC-seq) or DNase hypersensitivity assays. When integrated with GWAS using colocalization analyses^14^, caQTL datasets can help prioritize likely regulatory SNPs at the GWAS signals. However, these approaches remain constrained by study sample sizes and LD^15,16^. As a complementary approach to caQTL analyses, we and others have developed ATAC-seq allelic imbalance (ASAI) analysis to identify functional variants associated with diseases and traits^17–20^. Unlike population-level statistical associations from caQTL analysis, ASAI analysis directly assesses the allele-specific contribution of heterozygous SNPs to chromatin accessibility within individual samples. Because both alleles exist within the same cellular and environmental context, the intra-sample nature of ASAI reduces noise and increases signal. In addition, the short fragment sizes generated by ATAC-seq (typically less than 500 bp) can further enhance resolution for identifying functional candidates among linked SNPs.

In this study, we introduce an approach to nominate likely functional SNPs in the context of T2D using ASAI data. For complex diseases such as T2D, regulatory effects are often highly tissue-, cell type-, and developmental-stage specific^12,21–23^. We took advantage of the large amount of public deep ATAC-seq data from human T2D-relevant tissues, including skeletal muscle^15,16^, liver^24,25^, pancreatic islets^26–28^, adipose tissue^29,30^, and fluorescence-activated cell sorted (FACS) pancreatic islet endocrine cell populations^31^. This has enabled us to undertake a systematic ASAI analysis for T2D and related traits. We hypothesize that: (1) ATAC-seq peaks mark important regulatory elements within the genome; (2) T2D-associated SNPs within these regions harbor functional regulatory variants; (3) the imbalance of ATAC-seq reads at a heterozygous SNP provides an internally controlled measure of the functional importance of one allele over the other; and (4) the two alleles of a functional SNP are likely to be differentially associated with the accessibility of chromatin and TF binding in a relevant tissue. Under these assumptions, we systematically profiled ASAI across T2D disease-relevant tissues and identified strong candidate functional variants at multiple T2D GWAS loci. This approach presents a powerful strategy to further refine statistical fine-mapped credible set SNPs and can be applied to any disease or trait in tissues of interest to localize or identify the SNPs underlying GWAS signals.

## Results

### 1. Identification of ATAC-seq allelic imbalanced SNPs in multiple tissues

We assembled publicly available ATAC-seq raw sequence reads of 490 donors from multiple human tissues and cell types, including skeletal muscle (281 donors)^15,16^, liver (142 donors)^24,25^, pancreatic islets (24 donors)^26–28^, adipose tissue (14 donors)^29,30^, flow-sorted islet endocrine (alpha, beta and delta; five, four, and six donors, respectively) and exocrine cells (acinar and ductal; six and seven donors, respectively)^31^, and MEL1-derived beta cells (1 donor)^32^ (STable 1). We aligned reads to the reference genome (GRCh38) and performed reference allele mapping bias using WASP^33^. For each sample, we generated imputed genotypes using the TOPMed imputation reference panel^34^. We identified SNPs in donors with heterozygous genotypes and then generated the number of ATAC-seq read counts with either the reference or alternate alleles (see Methods; Figure 1A).

**Figure 1.**
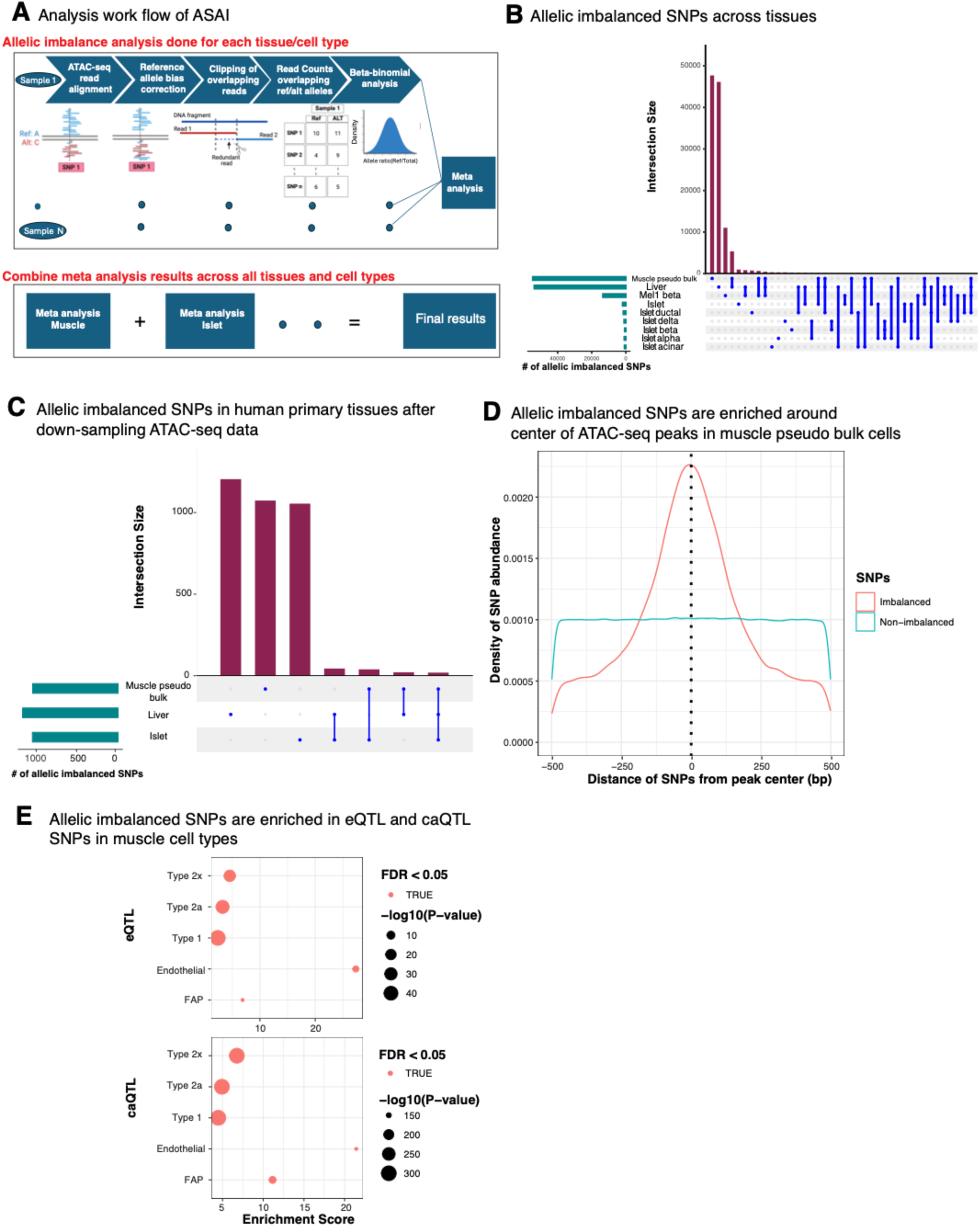
A. Analysis workflow of ATAC-seq reads for ASAI in SNPs across T2D disease-relevant tissues and cells. B. ATAC-seq allelic imbalanced SNPs across tissues and cells. “Muscle pseudo bulk” indicates pseudo bulk data generated by aggregating single-nucleus multiome ATAC-seq data across cell types. Note, we detected only 10 SNPs showing ASAI in adipose, so excluded it in the upset plot. See supplemental information for more details. C. ATAC-seq allelic imbalanced SNPs across human primary tissue samples using down-sampled ATAC-seq data. Down-sampling of ATAC-seq data was performed such that an equal number of donors with heterozygous genotypes and ATAC-seq reads were used to test ASAI of SNPs across the tissues. D. Allelic imbalanced SNPs are enriched around the center of ATAC-seq peaks in muscle pseudo bulk cells. E. ATAC-seq allelic imbalanced SNPs are enriched in eQTL (top) and caQTL SNPs (bottom panel) compared to all SNPs tested for ASAI in the muscle cell types.

In skeletal muscle, where we had access to single-nucleus multiome ATAC-seq data from 281 donors, we assessed ASAI in data aggregated across 13 cell types (“muscle pseudo bulk”) and aggregated in individual cell types^15^. We performed a beta binomial test to assess ASAI of ATAC-seq reads overlapping reference and alternate alleles per individual and then conducted a Z-score based meta-analysis across all samples for each SNP in a given tissue or cell type (Methods; Figure 1A). We tested 4,126,441 SNPs and identified a total of 119,949 SNPs that exhibited ASAI (FDR<0.05 for each tissue or cell type), the majority of which were observed in common muscle cell types and liver samples where the number of samples was substantially greater than in the other tissues (Figure 1B; Figure S1A-B; Supplemental data).

We next compared ASAI across tissues as variants associated with complex traits often exert their effects in trait-relevant tissues and cell types^35^. To account for differences in sample size and sequencing depth, we standardized the analyses for each SNP across the three primary tissues with abundant imbalanced SNPs (muscle, liver, and islet) by down-sampling both the number of heterozygous donors and sequencing coverage to the lowest levels observed across these tissues. We then repeated ASAI analysis on the down-sampled data for all SNPs identified as imbalanced in at least one tissue and applied multiple testing corrections in each tissue (see Methods). This yielded 3,454 SNPs with ASAI in at least one tissue. Among these, 3,330 SNPs were imbalanced in one tissue,106 SNPs were imbalanced in two different tissues, and 18 SNPs across all three tissues (Figure 1C), indicating that the majority of functional SNPs may contribute to regulation in only one of these T2D-relevant tissues. However, to take advantage of statistical power of tissues with large sample size and/or sequencing depth, we based all downstream analyses using the full data sets.

Imbalanced SNPs are enriched at the centers of ATAC-seq peaks compared to non-imbalanced SNPs (Figure 1D) and typically appeared as singleton imbalanced variants within peaks. As expected, ASAI SNPs were highly enriched among eQTL and caQTL SNPs in skeletal muscle common cell types ^15,16^ (Figure 1E), consistent with their role in regulating the transcriptomic and epigenetic profiles in these cell types.

### 2. ASAI analysis complements caQTL in nominating functional SNPs

An important question is whether ASAI provides additional functional information beyond that captured by caQTL analyses^25^. Traditional caQTL analyses are powerful in detecting population level changes in open chromatin regions due to genetic factors. However, caQTL signals often include multiple variants in high LD, making it difficult to pinpoint functional SNPs. In contrast, ASAI directly compares chromatin accessibility between alleles within the same heterozygous individual, providing a strong internal control that has been exposed to the same environmental and cellular conditions. For simplicity, we focus our comparisons on the lead caQTL SNPs within the corresponding ATAC-seq peaks (caQTL peaks) in each tissue. We compared SNPs showing ASAI with the lead caQTL SNPs located within the ATAC-seq peaks (see Methods) taking into account LD buddies in the same muscle cell types of Varshney *et al* (2024)^15,16^.

ATAC-seq peaks with ASAI SNPs are markedly more likely to be caQTL peaks than ATAC-seq peaks lacking ASAI SNPs (Hypergeometric P < 0.05 in major muscle cell types; Figure 2A). We calculated ASAI effect sizes (0.5 – Reference Ratio within ATAC reads)^36^ and found that the effect sizes of the lead caQTL SNPs under an ATAC peak are strongly correlated with those of ASAI when the SNPs also show ASAI (Pearson correlation r=0.92; P<2.2x10^-16^; Figure 2B, Type 1 muscle cells). Greater than 44.1% (3,546/8,045) of the lead SNPs identified by caQTL were also captured by ASAI (Figure 2C), showcasing the complementarity of the two methods to pinpoint likely functional candidates.

**Figure 2.**
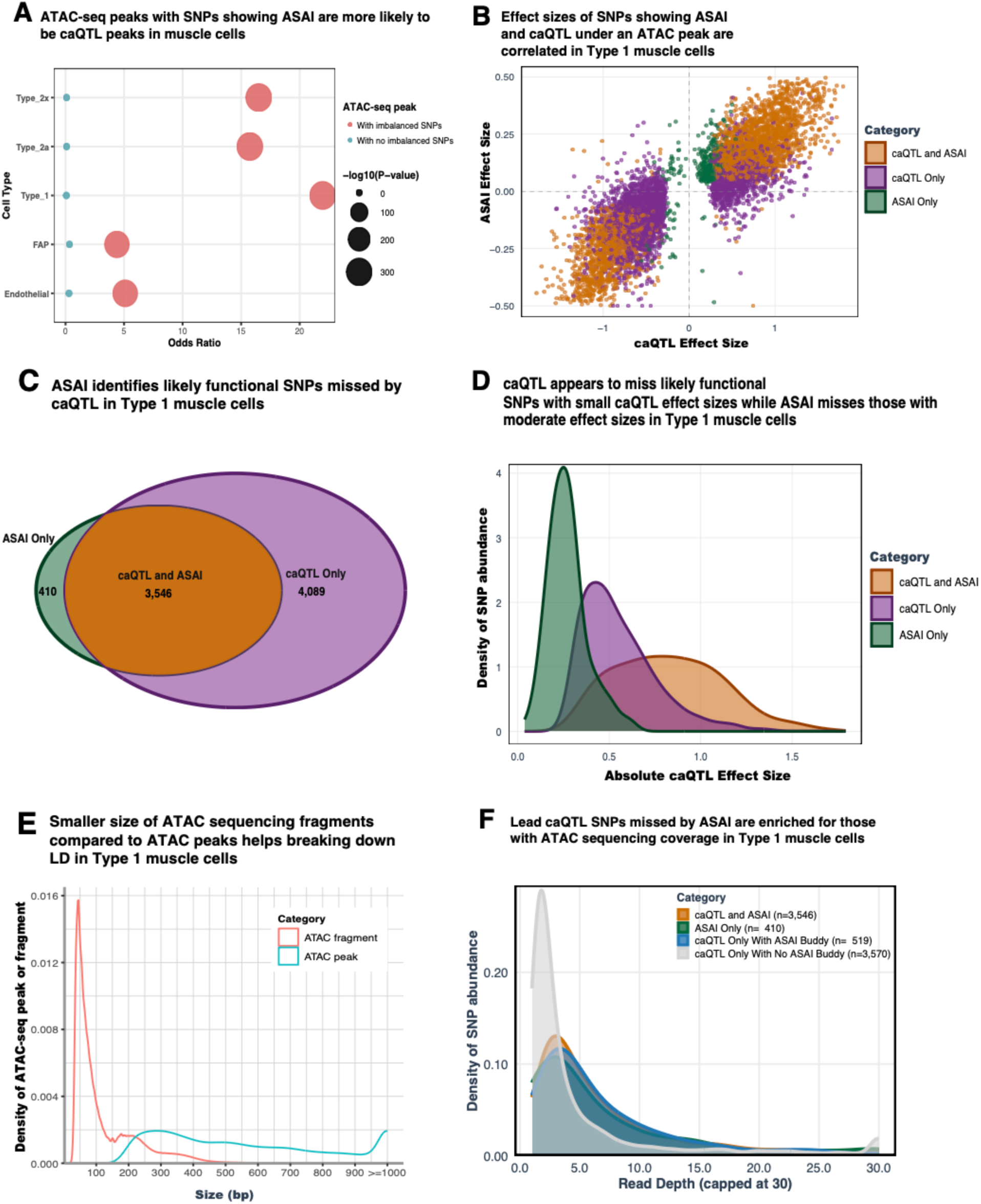
A. An ATAC-seq peak with imbalanced SNPs are more likely to be caQTL peaks compared to all ATAC-seq peaks in major muscle cell types. In contrast, peaks lacking an ASAI SNP are less likely to be a caQTL. P-value represents Hypergeometric P value of enrichment of ATAC-seq peaks with different categories of SNPs compared to all peaks in an individual muscle cell type. B. Effect sizes of SNPs in ASAI and caQTL are correlated in Type 1 muscle cells. C. ASAI identifies 410 likely functional SNPs missed by a caQTL analysis in Type 1 muscle cells, but caQTL calls thousands of signals not identified by ASAI. D. A caQTL appears to miss likely functional SNPs with moderate effect size in Type 1 muscle cells. E. Comparison of ATAC-seq fragment sizes and ATAC-seq peak widths in Type 1 muscle cells. ATAC-seq fragments are substantially smaller than the called peak regions, enabling ASAI to assess allele-specific contributions of each caQTL SNP in high LD regions at finer resolution. F. caQTL lead SNPs that are missed by ASAI belong to two categories: 1) 519 of 4,089 have another high LD SNP that is imbalanced in the same peak; 2) remaining 3,570 caQTL lead SNPs are less likely to be functional, and their nearby functional candidates are missed by ASAI at least partially due to scarce ATAC sequencing depth.

In addition to this shared set, we identified 410 SNPs showing ASAI (FDR < 0.05) that were not detected by caQTL analysis (“ASAI-only SNPs”; Figure 2C). These ASAI-only SNPs have smaller caQTL effect sizes than SNPs identified by both approaches (Figure 2D), suggesting that caQTL analyses may lack power to detect modest accessibility effects for some variants. For example, rs13051142 showed pronounced allelic imbalance in Type 1 muscle cells, with more ATAC-seq reads for the “A” allele than the “G” allele (aggregate reads mapped to “A” allele vs “G” at a proportion of 1,216/883; nominal P=1.6x10^-9^). However, its caQTL effect size was moderate (slope=0.23) and did not meet the FDR < 0.05 threshold. Notably, this SNP is also an eQTL in the same cell type^15,16^, where the “A” allele is associated with lower expression of *LCA5L* (Figure S2A). Together, this suggests that ASAI can identify likely functional variants with modest effects that may be missed by standard caQTL analyses with a relatively small sample size.

We also identified 4,089 caQTL lead SNPs within ATAC-seq peaks that did not meet our ASAI significance threshold (“caQTL-only SNPs”; Figure 2C). Among these, 519 SNPs (“caQTL-only with ASAI buddy”; 12.6%) have another linked SNP under the same peak that shows ASAI, suggesting that the lead caQTL SNP itself may not be functional, whereas the linked imbalanced SNP is (r²≥0.8). This improved resolution is a result of ASAI analysis to assess allele specific chromatin accessibility of nearby linked SNPs by taking advantage of the shorter length of ATAC-seq fragments relative to the broader span of ATAC-seq peaks (Figure 2E). The remaining 3,570 SNPs (“caQTL-only with no ASAI buddy”) include predominantly SNPs with lower ATAC sequencing depth, and to a lesser extent smaller minor allele frequency (MAF), compared to the SNPs identified by both approaches or by ASAI alone (Figure 2F; Figure S2B and S2C). This suggests that our study is underpowered to detect ASAI at variants with lower sequencing coverage or reduced representation.

### 3. ATAC-seq allelic imbalanced SNPs are enriched in transcription factor footprints in tissues relevant to T2D pathophysiology

Many transcription factors (TFs) require accessible chromatin to bind DNA and regulate target gene expression^37^. Pioneer TFs have the capacity to bind to target DNA sequences within compacted, “closed” chromatin and initiate opening of the regions for other conventional TFs to bind^38^. Therefore, SNPs exhibiting allele-specific effects on chromatin accessibility and modulating TF binding may point to functional regulatory variants. We scanned the ATAC-seq peak sequence regions using publicly available TF position weighted matrices (PWM) from the Cis-BP (v2) database^39^ and generated ATAC-seq footprint profiles accounting for the sequencing depth using CENTIPEDE^40^ on a per-sample basis.

We found that allelically imbalanced SNPs were strongly enriched within footprints for TFs expressed in the corresponding primary human muscle, liver, and islet tissues compared to all SNPs (Hypergeometric test; FDR < 0.05; Figure 3A). In contrast, no non-imbalanced SNPs under ATAC peaks showed enrichment in any tissue. Imbalanced SNPs were also more likely than non-imbalanced SNPs to disrupt conserved, high information-content nucleotides within TF motifs (Mann-Whitney test; P<2.2x10^-16^; Figure 3B), suggesting that imbalanced SNPs have higher impact on TF binding activity.

**Figure 3.**
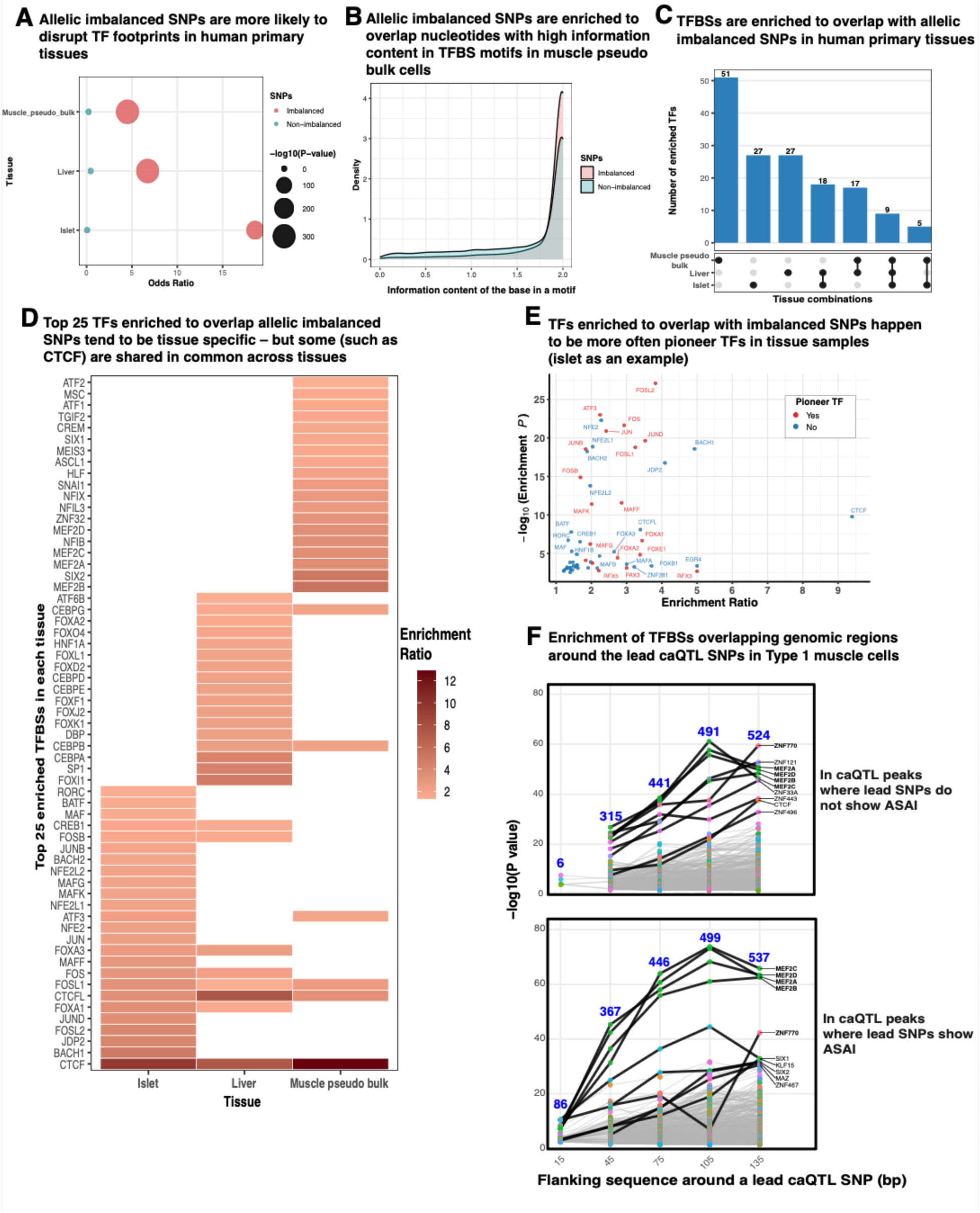
A. ATAC-seq allelic imbalanced SNPs are enriched in transcription factor (TF) ATAC-seq footprints compared to overall SNPs (imbalanced and non-imbalanced) in T2D relevant tissues. In contrast, non-imbalanced SNPs are not enriched in TF ATAC-seq footprints. B. ATAC-seq allelic imbalanced SNPs are enriched to overlap nucleotides with higher information content in TF footprints compared to non-imbalanced under ATAC peaks. X-axis indicates degree of information content of the nucleotides overlapping SNPs. 0 indicates a nucleotide in the motif is not conserved and 2 as highly conserved. C. Certain TFs are enriched to bind to regions with imbalanced SNPs in human primary tissues including muscle, liver and islet. The enrichment of some TFs tends to be stronger in one or two tissues, but others such as CTCF appear to be enriched across all three tissues. D. Top 25 TFs enriched to overlap ATAC-seq imbalanced SNPs in human primary tissues. E. TFs enriched that bind around imbalanced SNPs are more often pioneer factors compared to all TFs binding in any SNP regions. F. caQTL lead SNPs missed by ASAI (“caQTL Only With No ASAI Buddy”) are not strongly enriched to overlap with TFBSs, whereas nearby regions show increasing TFBS enrichment with larger flanking windows (top panel) similar to SNPs showing ASAI (bottom panel).

To determine which TF binding sites (TFBSs) were disrupted, we examined TF motifs within 15 bp of imbalanced and non-imbalanced SNPs located in ATAC-seq peaks across tissues (see Methods). We found that imbalanced SNPs showed strong enrichment for TFs important in each tissue, in muscle: members of the myocyte enhancer factor family (MEF2A, MEF2B, MEF2C, MEF2D), musculin (MSC), and myoblast determination protein 1 (MYOD1) (Figure 3C-D, Figure S3A and D); in liver: CCAAT/enhancer binding protein α (CEBPA), hepatocyte nuclear factors (HNF4A and HNF4G) (Figure S3B); and in islets: members of the Jun transcription factor family (c-Jun, JunD, JunB), regulatory factor X (RFX), Maf transcription factor A and B (MAFA and MAFB) (Figure S3C). We also observed enrichment of several housekeeping TFs, including CTCF and FOSL1 across all three tissues (Figure 3C-D, Figure S3D). Notably, we observed an over representation of pioneer factors among these TFs in all three tissues (Hypergeometric test, FDR<0.05) suggesting that imbalanced SNPs may modulate master regulators of differentiation and development of these respective tissues (Figure 3E; STable 4).

We further investigated the nature of the 3,570 “caQTL-only with no ASAI buddy” (Figure 2F) by testing for enrichment of TFBSs overlapping these SNPs. Focusing only on a 15 bp window around the lead SNPs in these caQTL-only peaks, we noted a much lower enrichment for muscle-relevant TFBSs in type 1 muscle cells than with ASAI SNPs (six vs. 86 in top vs bottom panel, Figure 3F). However, increasing the flanking windows around “caQTL only” lead SNPs showed enrichment for these same muscle-specific TFs, suggesting that nearby linked variants may represent the true functional candidates, flagged by caQTL analysis because of the longer length of ATAC peaks compared to sequencing reads (Figure 2E). These neighboring variants may have been missed by ASAI due to limited sequencing depth and sample size.

### 4. ASAI analysis helps to nominate likely functional SNPs for T2D and related traits

An ASAI analysis provides a less biased way to assess the allele-specific effects of individual SNPs on chromatin accessibility and *cis*-regulatory activity. To test if this approach captures likely functional variants in the context of GWAS, we intersected SNPs tested for ASAI with 99% credible sets for 338 T2D association signals where the credible sets have been published^1^. We found that SNPs showing ASAI were significantly enriched in the T2D credible sets for muscle pseudo bulk cells (Hypergeometric test, P=9.01x10^-10^; enrichment score 1.77) and for other related traits (Figure S4A-C), indicating the validity of ASAI for identifying candidate functional variants.

We next tested whether ASAI analysis could refine GWAS statistical fine-mapping by reducing the number of candidate variants within credible sets. Of the 8,351 SNPs within the 99% credible sets across 338 GWAS T2D association signals^1^, we identified 256 SNPs from 123 signals that showed ASAI in at least one tissue/cell type (Figure 4A and 4B; Figure S4D). 119 out of 123 signals have more than one SNP in the credible set and the ASAI refinement substantially reduced the number of predicted functional SNPs for experimental follow-up (Figure 4C). Notably, 71 of the 119 signals contained exactly one imbalanced SNP, while the remaining 48 signals harbored multiple imbalanced variants, often across distinct tissues or regulatory elements. For instance, at the *CDKN2A/B* locus, we identified two ASAI SNPs: rs10811647 in Type 1 muscle cells (T2D risk allele “G” vs. non-risk “C” at a proportion of 351/268; P=1.59x10^-5^) and rs6475604, located 12 kb upstream, in liver tissue (T2D risk allele “C” vs. non-risk “T” at a proportion of 144/80; P=5.18x10^-7^) (STable 2). Furthermore, in muscle cell types with the largest sample size and thus power, imbalanced T2D credible SNPs overlapped with TF footprints more often than non-imbalanced SNPs in muscle pseudo bulk, Type 1, 2a and 2x cells (FDF<0.05), supporting their likely functional roles (Figure 4D).

**Figure 4.**
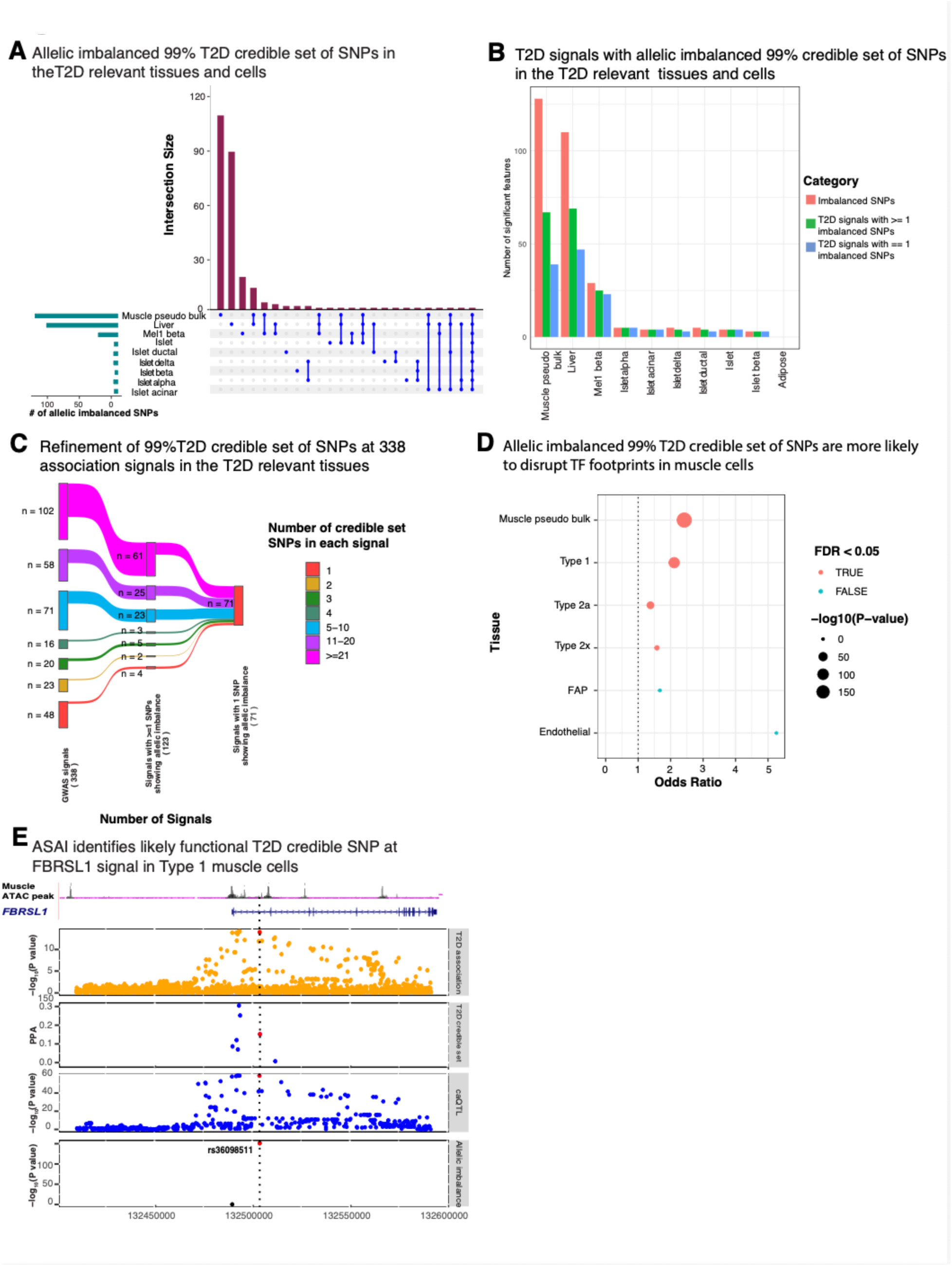
A. Allelic imbalanced 99% T2D credible set of SNPs in the T2D relevant tissues and cells. B. T2D signals with allelic imbalanced 99% credible set of SNPs in the T2D relevant tissues and cells. C. Refinement of 99% T2D credible set of SNPs at 338 association signals across T2D disease related tissues and cells. D. Allelic imbalanced 99% credible set of SNPs are likely to disrupt TF footprints compared to overall credible set of SNPs in muscle pseudo bulk, as well as major cell types. E. ASAI analysis nominates a likely functional T2D credible SNP (rs36098511) at the *FBRSL1* signal in Type 1 muscle cells. Panels from top to bottom: (1) -log_10_(T2D association P value) based on ^1^ study. (2) Posterior probability of T2D association for 99% credible set of SNPs. (3) -log_10_(caQTL nominal P value) for the caQTL peak overlapping rs36098511. (4) -log_10_(ASAI P value).

We highlight the T2D association signal near *FBRSL1*, which also overlaps a blood glucose GWAS signal^41^ to illustrate the use of ASAI to nominate a likely functional variant from a pool of multiple candidate variants. Mahajan *et al.* (2022) statistically fine-mapped the T2D association signal to a 99% credible set of seven SNPs, including rs36098511 (posterior probability of association, PPA=0.15)^1^, which overlap non-coding intronic regions within the *FBRSL1* gene (Figure 4E). Two of these seven SNPs overlapped an ATAC-seq peak in muscle pseudo bulk cells and were in strong LD with the lead GWAS variant rs12811407 (r^2^≥0.8)^1^. Varshney *et al.* (2024) colocalized a caQTL peak overlapping rs36098511 with the T2D signal in muscle Type 1, Type 2a, and Type 2x cells^15,16^. In our analysis, only rs36098511 displays significant ASAI in muscle pseudo bulk and common muscle cell types, with the non-risk “A” allele having substantially more ATAC reads than the risk “T” allele (aggregate reads mapped to “A” allele over “T” allele at a proportion of 1,838/259 across donors with heterozygous genotypes; P=6.94x10^-156^. In contrast, the second candidate SNP rs77773608 shows no imbalance (“C/A” = 1,708/1,754). The ATAC-seq peak overlapping rs36098511 is only present in muscle pseudo-bulk or common cell types, but not in islet or liver, suggesting that rs36098511 may affect glucose metabolism through skeletal muscle-specific regulatory activity.

Next, we extended this approach to additional T2D-related traits. We identified 1,175 SNPs in 99% credible sets that also showed imbalance for 569 association signals for HbA1C, 290 SNPs in 153 signals for blood glucose^41^, and 130 SNPs in 51 signals for random glucose^42^ (Table 1; Figure S4E-G). Similar to the T2D analyses, ASAI substantially narrowed the number of candidate functional SNPs within GWAS credible sets and prioritized a smaller number of SNPs for future experimental validation.

**Table 1.** Refinement of T2D and related traits GWAS 99% credible set of SNPs for likely functional variants using ASAI analysis in disease relevant tissues and cells.

| Trait | Number of association signals | Number of 99% credible SNPs | Number of association signals where 99% credible SNPs tested | Number of ATAC-seq imbalanced SNPs | Number of association signals with ATAC-seq imbalanced SNPs | Number of 99% credible set of SNPs in signals with imbalanced SNPs | % of association signals with ATAC-seq imbalanced SNPs | Number of association signals with only one ATAC-seq imbalanced SNPs | % of association signals with one ATAC-seq imbalanced SNPs | % of imbalanced SNPs out of 99% credible set SNPs in signals with imbalanced SNPs |
| --- | --- | --- | --- | --- | --- | --- | --- | --- | --- | --- |
| T2D | 338 | 8,581 | 314 | 256 | 123 | 5,044 | 36.39 | 71 | 21.01 | 5.08 |
| Random glucose | 133 | 3,445 | 125 | 130 | 51 | 2,243 | 38.35 | 25 | 18.80 | 5.80 |
| HbA1C | 1,329 | 52,285 | 1,232 | 1,175 | 569 | 39,436 | 42.81 | 254 | 19.11 | 2.98 |
| Blood glucose | 313 | 12,785 | 297 | 290 | 153 | 10,184 | 48.88 | 79 | 25.24 | 2.85 |

### 5. Experimental validation of ATAC-seq imbalanced T2D SNPs

As one means of validating ASAI, we surveyed the literature and compiled a list of T2D SNPs with prior experimental evidence for allele-specific regulatory activity using one or more functional assays such as MPRA, luciferase assays, EMSAs, ASAI in regulatory elements (FDR<0.05) (ATAC-seq, ChIP-seq), or CRISPR editing approaches (Methods). Among SNPs within the 99% credible sets for 338 T2D association signals, we found prior experimental evidence supporting allele-specific functionality for 109 SNPs across 63 signals (STable 2 and 3; Figure 5A). Of these 109 SNPs, 23 SNPs across 21 signals also exhibited significant ASAI in the tissues analyzed in our study (STable 2). These findings provide validation for the ASAI framework and further support the causality of these variants. Some of these 23 SNPs are imbalanced in only one tissue or cell type, such as rs4841132 at the *MSRA-XKR6* locus in liver, whereas others, like rs7163757 at the *C2CD4A/B* locus, show strong ASAI in multiple tissues, including islet acinar cells, liver, as well as abundant muscle cells (Figure S5A).

**Figure 5.**
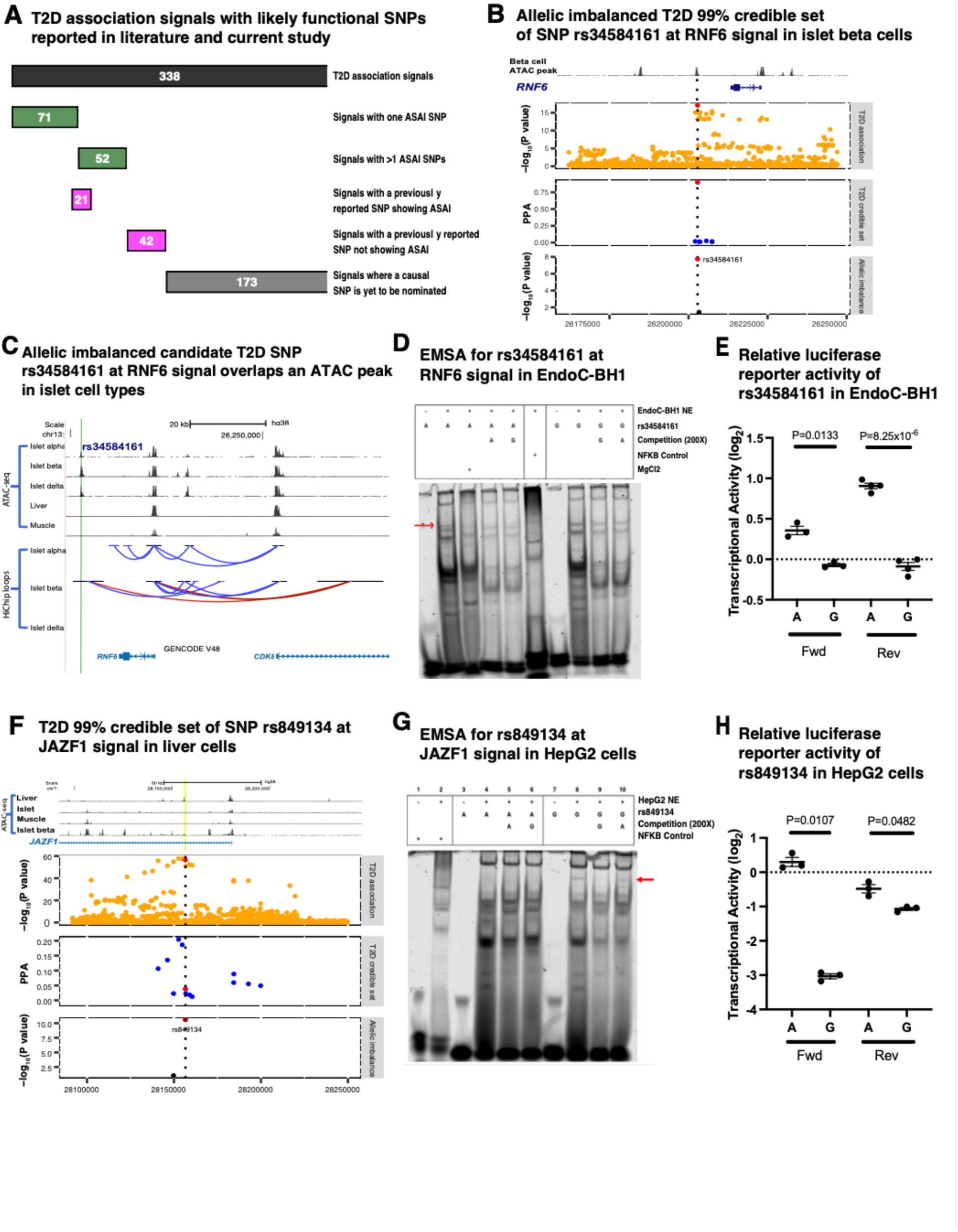
A. T2D association signals with previously reported functional SNPs. Top black bar: total number of T2D association signals and 99% credible set of SNPs assessed; two green bars: association signals with ATAC-seq imbalanced SNPs in current study; two magenta bars: signals with previously reported functional SNPs; bottom grey bar: signals with no previously reported functional SNPs nor ATAC-seq imbalanced SNPs. B. T2D SNP rs34584161 at RNF6 signal in islet beta cells. Panels from top to bottom: (1) ATAC-seq peak in islet beta cells. (2) -log_10_(T2D association P value). (3) PPA of T2D GWAS analysis. (4) -log_10_(ASAI P value). C. The location of rs34584161 is indicated by a vertical light green line. This overlaps ATAC-seq peaks present only in FACS islet endocrine cells: alpha, beta and delta cells. HiChip data show that the region overlapping this SNP is in close physical proximity with *CDK8* and *RNF6*. D. Electromobility shift assays probing T2D risk and non-risk alleles of rs34584161 at the *RNF6* signal using EndoC nuclear extracts. Red arrow indicates a band only present in well with T2D risk “A” allele and competed away by unlabeled “A”, indicating that “A” allele is being bound by an unknown protein. E. The 201-bp sequences encompassing rs34584161 containing either the “A” or “G” allele were cloned upstream of a minimal promoter luciferase reporter in both forward (Fwd) and reverse (Rev) orientation and transfected into the human beta cell line EndoC-BH1. Luciferase activity was normalized and plotted on a log₂ scale relative to control. Risk allele “A” showed significantly greater transcriptional activity than the non-risk “G” allele. Points represent independent biological replicates; bars indicate mean±SEM. Dotted line denotes activity of the empty vector control. Statistical significance was assessed using a two-sided Welch’s corrected unpaired t-test (P = 0.0133). F. Nominating SNP rs849134 at the *JAZF1* signal in liver cells. Panels from top to bottom: (1) ATAC-seq peaks in liver, islet, muscle and islet beta cells. (2) -log_10_(T2D association P value). (3) PPA of T2D GWAS analysis. (4) -log_10_(ASAI P value). G. Electromobility shift assays probing T2D risk and non-risk alleles of rs849134 at the *JAZF1* signal using HepG2 nuclear extracts. Red arrow indicates a band only present in well with T2D non-risk “G” allele and competed away by unlabeled “G”, indicating that “G” allele is being bound by an unknown protein. H. The 201-bp sequences encompassing rs849134 containing either the “A” or “G” allele were cloned upstream of a minimal promoter luciferase reporter in both forward (Fwd) and reverse (Rev) orientation and transfected into the human liver line HepG2. Luciferase activity was normalized and plotted on a log₂ scale relative to control. T2D non-risk allele “G” showed lower transcriptional activity than the risk “A” allele. Points represent independent biological replicates; bars indicate mean±SEM. Dotted line denotes activity of the empty vector control. Statistical significance was assessed using a two-sided Welch’s corrected unpaired t-test (P=0.0107).

We next experimentally evaluated two additional T2D GWAS loci, *RNF6* and *JAZF1*^1^. At the *RNF6* locus, seven SNPs comprise the 99% credible set for the T2D association signal (Figure 5B). Among these, only rs34584161 showed significant ASAI, with more ATAC-seq reads mapping to the T2D-risk “A” allele compared to the non-risk “G” allele (Figure S5B) in sorted human pancreatic islet alpha, beta, and delta cells. This SNP overlaps a strong ATAC-peak in islet cells but not in muscle or liver tissue (aggregate reads mapped to “A” allele over “G” allele at a proportion of 222/19 across donors with heterozygous genotypes in islet beta cells; P=1.75x10^-8^; Figure 5C; Figure S5C). No ASAI was detected (FDR>0.05) for the remaining six credible-set SNPs.

Although some experimental characterization has been performed for the *RNF6* locus, none has been conducted in the immortalized human pancreatic beta cell line EndoC-BH1. We performed EMSAs on all seven SNPs in the 99% credible set for this signal using nuclear extracts from EndoC-BH1 cells. Of these, only rs34584161 exhibited allele-specific protein-DNA binding, with the risk “A” allele forming an additional protein-DNA complex (Figure 5D). ATAC-seq TF footprint analysis suggested that the G allele of this SNP may disrupt binding of Maf, Fos, and Jun family proteins, though supershift assays did not support direct binding of MafA, MafF, MafG, and MafK. We next tested the effect of rs34584161 on transcriptional activity using luciferase reporter assays in EndoC-BH1 cells. The risk “A” allele showed significantly greater transcriptional activity than the non-risk “G” allele in both forward (P=0.013) and reverse orientations (Welch’s two-sided t-test, P=8.25x10^-6^; Figure 5E). Previous studies identified rs34584161 as the lead islet eQTL SNP for two nearby genes *RNF6* and *CDK8* in human islets, with the risk allele associated with increased expression of both genes^43^. HiChIP data further show chromatin interactions of the region overlapping rs34584161, the first intron of *CDK8,* and the *RNF6* promoter specifically in islet beta cells^31^ (Figure 5C). Together, these findings support rs34584161 as a likely functional variant at the *RNF6* locus that affects the transcriptional regulation of target genes by modulating the binding of an unknown activator TF.

At the *JAZF*1 locus, two SNPs (rs849134 and rs849135) out of 13 in the 99% credible set for its T2D association signal overlap a liver-specific ATAC-seq peak. rs849134 showed one of the strongest ASAI signals in liver (aggregate reads mapped to T2D non-risk “G” allele over risk allele “A” at a proportion of 1,005/221 across donors with heterozygous genotypes; P=2.77x10^-71^; Figure 5F; Figure S5D) and is also the lead caQTL SNP in a liver study^24^. In contrast, rs849135 shows modest imbalance while lying 191 bp away from rs849134 (aggregate reads mapped to “A” allele over “G” allele at a proportion of 248/170; P=3.43x10^-4^, adjusted P=0.02). We performed EMSAs on both SNPs using HepG2 and EndoC-BH1 nuclear extracts.

Comparing the SNPs, only rs849134 exhibited allele-specific protein-DNA binding, where the non-risk “G” allele formed an additional complex specifically in conjunction with HepG2 nuclear extracts (Figure 5G; Figure S5E). Luciferase assays in HepG2 cells further showed reduced transcriptional activity of the non-risk “G” allele in both forward (Welch’s two-sided t-test, P=0.011) and reverse orientations (P=0.048; Figure 5H), suggesting that rs849134 alters the binding of a liver-specific repressive TF. Consistent with the EMSA results, we did not observe differential transcriptional activity for rs849135 (forward orientation P=0.16, reverse orientation P=0.71; Figure S5E). Together, these findings nominate rs849134 as a likely functional regulatory variant at the *JAZF1* locus contributing to T2D risk through liver-specific regulatory mechanisms while rs849135 is a linked variant in high LD with rs849134.

Finally, we asked whether ASAI could identify functional variants that were missed by statistical fine-mapping of credible sets. We assembled a list of SNPs in LD with the lead T2D GWAS SNPs from^1^ (r² ≥ 0.8; UKBB European panel) and intersected them with ASAI-positive SNPs identified in this study. We identified 24 additional T2D signals (7% of 338 signals) containing imbalanced LD SNPs outside the published 99% credible sets (STable 5). For example, none of the published nine 99% credible SNPs at the *SLC2A2* locus showed significant ASAI. However, we identified a nearby non-synonymous coding SNP, rs5398, with strong allelic imbalance in liver (read counts for T2D risk allele “G” vs. non-risk “A” at a proportion of 973/666; P=1.35×10⁻⁸) (Figure S5F). This suggests rs5398, which is in strong LD (r² = 0.98) with the lead GWAS SNP rs8192675, is a potential overlooked functional variant that affects both chromatin access and coding potential, located within the gene *SLC2A2* that encodes the GLUT2 glucose transporter^44^.

## Discussion

Resolving the molecular processes that mediate genetic risk for T2D remains a challenge because most disease-associated variants are located in non-coding regions within tight LD blocks, and functional characterization of these signals requires knowledge of the specific tissues and cell types in which they operate^2,45^. To our knowledge, this is the largest study to leverage ASAI to refine the likely causality of T2D GWAS SNPs in disease-relevant tissues, nominating functional variants for 123 of 338 GWAS T2D association signals^1^. The reliability of the findings was supported by comparison with prior functional studies and by experimental validation of proposed causative SNPs at *RNF6* and *JAZF1*.

Our findings suggest that the majority of imbalanced SNPs show tissue specificity and are more likely to disrupt critical transcription factor binding sites (TFBSs) compared to non-imbalanced SNPs. These variants often disrupt key TFBSs relevant to the function of the tissue, as shown here for muscle, liver, and islets. These TFs are more frequently pioneer factors that facilitate open chromatin accessibility and shape the epigenetic landscape of cells^38^. These observations highlight the importance of investigating SNP function in the appropriate biological context. However, it is also important to note that smaller sample size and/or lower sequencing depth in some tissues (for example, adipose) may have contributed to a reduced ability to identify imbalanced SNP.

Our results also argue that multiple functional variants can contribute to disease susceptibility at a single GWAS locus^46,47^. For example, at the *CDKN2A/B* locus, we identified two ASAI SNPs: rs10811647 in Type 1 muscle cells and rs6475604 in liver tissue. This suggests that multiple distinct regulatory elements overlapping different SNPs may contribute to T2D risk in a tissue-dependent manner within the same locus. Importantly, we must recognize two possibilities that could result in ASAI for multiple SNPs at a locus: coordinated regulation of distant regulatory elements that harbor the SNPs^24^, and high LD between SNPs in close proximity as demonstrated at the *JAZF1* locus in our study.

We did not detect SNPs with ASAI in 42 of the 63 signals with previously reported functional SNPs, nor in 173 of the 338 T2D GWAS loci (Figure 5A). The ASAI approach would only capture SNPs that affect accessible chromatin regions in the tissues sampled for analysis. Several factors may explain the absence of ASAI SNPs for these 42 signals: (1) the power to detect imbalance is constrained by sample size and heterozygosity, especially in this study for adipose, islets, and islet cell subtypes (Figure S2B); (2) some causative SNPs reside in transcripts and exert their effects through mechanisms that do not impact chromatin accessibility, such as altering coding sequences (e.g., rs13266634 in *SLC30A8*)^48^, splice sites, or miRNA binding (e.g., rs2229295 in the 3′ UTR of *HNF1B*, which modulates T2D risk through mediating the binding of miRNAs hsa-miR-214-5p and hsa-miR-550a-5p)^49^; (3) some SNPs may be functional only in tissues, developmental stages, or contexts not profiled within our study; (4) some functional SNPs influence regulatory activity without altering chromatin accessibility. For example, rs7132908 at the *FAIM2* locus does not show allelic imbalance in our study, but it was reported to affect the strength of an enhancer in Mel1 iPSC-derived beta cells^32^ (STable 3); and (5) statistical fine-mapping may have failed to capture the true causal SNPs within the 99% credible sets, as we have shown in the case of the signal at the *SLC2A2* locus.

The accuracy to detect allelic imbalance depends on genotype quality^36,50^. To assess this in skeletal muscle, we used data from whole genome sequencing of 281 samples and confirmed high concordance of imputed genotypes for the imbalanced SNPs (99.5% for all sites; 99.1% for heterozygous sites), indicating that ASAI analysis is unlikely to be significantly confounded by underlying genotype quality as long as standard protocols are used. We also used WASP to correct reference allele mapping bias, which improved detection power by increasing alternate allele representation (mean 0.488 vs. 0.478; Wilcoxon P=6.4×10⁻¹⁰).

We also sought to systematically compare caQTL and ASAI approaches with this data set and found them to be highly complementary approaches with distinct strengths and weaknesses. caQTL uses population-level data from all three genotypes at a variant to identify regulatory regions and enables quantitative colocalization analyses with GWAS signals. However, caQTL approaches generally require larger sample sizes. In contrast, ASAI analyzes only heterozygous individuals and thus benefits from an internal control where both alleles are assessed within the same cellular and environmental context, making it valuable in settings with more limited sample sizes, as long as the minor allele frequency is not too low. In our study, the two approaches showed substantial overlap in their nomination of likely functional SNPs, while each also identified candidate functional SNPs missed by the other. caQTL analyses appeared to miss SNPs with modest effect sizes, while ASAI was more limited by sequencing depth. Importantly, ASAI can reduce the number of likely candidates when multiple closely linked caQTL SNPs are present within the same ATAC-seq peak. This increased resolution is likely due to ASAI measuring allele-specific chromatin accessibility at the level of smaller ATAC-seq fragments than the larger peak window traditionally used for caQTLs. Consistent with this idea, recent work showed that focusing on the smaller ATAC-seq fragments in nucleosome free regions provides higher resolution for nominating functional variants^51^.

Taken together, our results indicate the value of integrating both caQTL and ASAI analyses to improve power for signal detection and functional variant prioritization. In the context of GWAS signals, the strongest evidence for a functional variant emerges when caQTL and ASAI are applied together: (1) a caQTL colocalizes with a GWAS signal, supporting a shared likely functional variant; (2) the candidate variant lies directly within the accessible chromatin peak, refining the most likely functional variant; and (3) the variant exhibits allelic imbalance, validating the molecular mechanism at the allele in a controlled individual. In summary, we find ASAI analysis to be a powerful tool that can substantially refine the 99% credible set of GWAS SNPs in T2D, HbA1C, and blood glucose traits for more than 18% of the signals (Table 1), allowing for substantially improved identification of likely functional variants that contribute to the disease or trait susceptibility. These SNPs tend to disrupt TFBSs in a tissue-specific way. The data generated from this study should be a valuable resource for studying the functional genomics of T2D and other relevant traits. By increasing the sample size and sequencing depth, future analyses can help toward more efficient and precise identification of likely functional SNPs. A similar approach can be applied to other common disorders where genotyping and ATAC-seq data can be made available from disease-relevant tissues.

## Data, Materials, and Software Availability

Genome sequencing data of skeletal muscle samples were deposited to dbGaP (phs001579 and phs001048). Full summary statistics of all tested SNPs in tissue/cell type have been deposited to the Common Metabolic Diseases Knowledge Portal (https://t2d.hugeamp.org/downloads.html). Software programs used to perform beta-binomial test and subsequent meta-analysis are available at: https://github.com/NHGRI/MGS/tree/master/ATAC-seq_allelic_imbalance.

## Data Availability

All data produced in the present work are contained in the manuscript.

https://t2d.hugeamp.org/downloads.html

## Acknowledgements

The authors thank the individuals and their families who participated in this research study. The authors extend their gratitude to collaborators of the FUSION project (The Finland-United States Investigation of NIDDM) for collecting molecular and clinical data for the participants. This research was supported [in part] by the Intramural Research Program of the National Institutes of Health (NIH) through ZIA HG000024, ZIA BC011798, R01 DK072193, DK062370, and RC2 DK144819. CNS was supported by the American Diabetes Association (#11-22-JDFPM-06). The contributions of the NIH author(s) are considered Works of the United States Government. Subsets of the samples/data used for the research were obtained from THL Biobank (study number THLBB2025_5). The findings and conclusions presented in this paper are those of the authors and do not necessarily reflect the views of the NIH or the U.S. Department of Health and Human Services.

## Author contributions

NN, DLT, CCR, HT, MB, LLB, MEE, KLM, SCJP, and FSC designed research; NN, HXL, CJMR, AV, AJS, CCR, DLT, HT, AB, BNL, DX, MB, EA, SC, LGB, and FSC performed research; HXL, CJMR, AJS, DX, AB, EA, SC, LLB, and MEE contributed new reagents/analytic tools and experimental work; NN, NS, TY, HXL, CJMR, CCR, DLT, HT, AV, KWC, BNL, HS, KAB, and EW analyzed the data; and NN, HXL, CJMR, AV, KLM, SCJP, and FSC wrote the paper. All authors reviewed and commented on the manuscript.

## Methods and materials

### 1. ATAC-seq data preparation and peak calling

We collected ATAC-seq raw sequence files of multiple human tissues and cell types, including liver (142 donors) ^24,25^, pancreatic islet (24 donors) ^26–28^, adipose (14 donors) ^29,30^, FACS pancreatic islet endocrine cells (alpha, beta, and delta with five, four, and six donors respectively) and exocrine cells (acinar and ductal with six, and seven donors, respectively) ^31^, and MEL1 iPSC derived beta cells ^32^. We only included donor data with paired-end sequence reads. We excluded any duplicate set when two studies used the same data set. For details of samples included in each cell tissue/cell type, see STable 1.

We obtained cell-type specific aligned bam file (GRCh38 reference genome) and an aggregated one for all cells (“muscle pseudo bulk”). for each of 281 muscle samples ^15,16^. For all tissue samples, we consistently reprocessed fastq files, aligned reads, and called peaks using the same approach described in the Varshney et al (2024) without subsampling reads ^15,16^. Briefly, we trimmed adaptor sequences using CTA (v0.1.2) (https://github.com/ParkerLab/cta) and aligned the trimmed reads to the GRCh38 genome assembly using ‘BWA-MEM’ (v0.7.17-r1194)^52^. We removed duplicate reads with ‘GATK MarkDuplicate’ (v4.1.9.0), filtered for autosomal, properly paired reads with mapping quality ≥30 with ‘samtools’ v1.9 ^53^, and retained uniquely aligned primary reads per library/donor. We removed one copy of overlapping read pairs for short fragments using “clipOverlap” of bamUtil (https://genome.sph.umich.edu/wiki/BamUtil). We converted the filtered BAM files to BED files using the ‘bamtobed’ function from ‘bedtools’ (v2.26.0) ^54^ and called narrow peaks using ‘MACS2’ (v2.2.7.1) ^55^. We removed candidate peaks that overlap with ENCODE blacklists ^56^ and controlled for a false discovery rate (FDR) of 0.05.

For each tissue/cell type, we merged ATAC peaks across all donors and generated a master set of peaks for the tissue/cell type for downstream analyses.

### 2. SNP chip genotype preparation and missing genotype imputation

We genotyped DNA of 24 pancreatic islets using Infinium Omni2.5Exome-8 BeadChip array v1.3 (Illumina, San Diego, CA) at the NHGRI Genomics Core facility, resulting in a call rate of 99.7% (out of 2,612,357 SNPs). We mapped the array probe sequences to the GRCh37 (hg19) genome assembly using novoalign v2.07.11 (http://www.novocraft.com/products/novoalign) and filtered variants with ambiguous probe alignments and allele frequency discordant with the 1000G phase 3 release panel as previously described ^32^. After filtering the genotypes, we used the remaining 1,589,371 SNPs for genotype imputation on the Michigan TOPmed Server (Minimac v4) ^34^. Similarly, we imputed genotypes using 2.5M exome SNP chip data of the remaining donors. Finally, we generated imputed genotypes in the GRCh38 reference panel for all SNPs (r² >0.3) included in the TOPmed panel and used the genotypes for downstream analyses.

For remaining samples, we obtained Exome2.5M SNP array genotypes from the respective study and imputed genotypes using the same TopMed reference imputation panel ^34^.

### 3. Comparison of TopMed Imputed genotypes against variants called using WGS

We performed the WGS of 281 FUSION muscle samples and called variants ^57^. To ensure imputed genotypes were of high quality, we assessed imputation quality of the SNPs tested for ATAC-seq allelic imbalance using variants called on the genome sequence data of the same samples. We used imputed genotypes as the reference and assessed SNPs with good genotype quality scores in the GATK calls (GT≥10) in 80% of samples (out of 281 samples) and the ones with MAF > 0.01. We calculated the percentage of genotype concordance between two approaches and generated a proportion of overall SNPs with matching genotypes and that of heterozygous genotypes.

### 4. ATAC-seq allelic imbalance analysis

We performed ATAC-seq allelic imbalance analysis for each sample using the aligned reads described in *ATAC-seq data preparation and peak calling).* We applied ‘WASP’ (v0.3.4) ^33^ to quantify read counts while controlling for reference allele mapping bias at heterozygous variants in each sample using imputed genotypes with an imputation quality r^2^>0.3. We took two different approaches to perform allelic imbalance analyses: 1) Using the ATAC-seq read counts mapped to reference and alternate alleles of each SNP, we performed a two-sided beta-binomial test (modified ‘binom.test’ function in R v4.2.2) in each donor. Since we are interested in assessing accumulative effect for each SNP, we performed a meta-analysis across donors with ≥1 total reads using Stourffer’s Z-score method ^58^ ^59^, weighting the Z-score from each donor by the total read counts overlapping the SNP (‘sumz’ method from ‘metap’ R package v1.8). This process generated a meta analysis P value per SNP. 2) Across all donors with heterozygous genotype for each SNP, we summed the number of reads aligned to the reference and alternate alleles and constructed “pseudo bulk” read counts for two alleles. Next, we performed the previously mentioned beta-bionomial test and generated one P value per SNP. Finally, for both approaches, we controlled for the number of tests using the Benjamini-Hochberg procedure ^60^ across all tested SNPs in each tissue/cell type effect analysis. We considered SNPs to be ATAC-seq imbalanced at FDR<0.05 for a given tissue/cell type and generated a set of study-wide imbalanced SNPs per tissue/cell type. Two approaches resulted in highly correlated test statistics, and we reported results from the first approach in the paper.

### 5. Comparison of caQTL and ASAI

We downloaded muscle cell type specific caQTL permutation test results for the lead caQTL SNPs and their respective 300bp ATAC-seq summits (https://zenodo.org/records/16573423). The caQTL analysis measured ATAC-seq read counts in a 301bp summit region of a peak. Therefore, a SNP under an ATAC peak but outside the summit region still can be tested for ASAI. To accommodate these SNPs, we intersected the summits tested in the caQTL analysis with the narrow peaks called within this study (See section *ATAC-seq data preparation and peak calling)*. We assigned the narrow peak with the largest overlap with an ATAC summit to be the corresponding ATAC peak for the summit in the permutation test (lead SNP and ATAC-summit). We used permutation test results of Type 1 muscle cells as an example and presented the results in the manuscript.

Next, we focused on the caQTL lead SNP-narrow peak pairs where the lead SNP is under the tested ATAC-narrow peak (aka lead SNP ATAC-summit pair) and that were also assessed for ASAI. We identified LD buddy SNPs of the caQTL lead SNPs using plink (v1.90b6.21) for 281 donors that we have imputed genotypes. We restricted LD buddies of a lead SNP to be under the same peak as the lead SNP and with high LD (r²≥0.8). We considered such LD buddies of a lead caQTL SNP to be additional caQTL SNPs for simplicity (Figure 2F).

### 6. Identification of TF ATAC-seq footprints

To assess the effect of SNPs on regulatory elements, we performed a TF footprint analysis in the merged ATAC-seq peaks for per donor and tissue/cell type individually as described ^32^.

Briefly, we generated a fasta file containing two sequences for each ATAC-seq peak, one for each allele of overlapping SNPs. We scanned these sequences for position weight matrices (PWMs) of the directly determined TF motifs included in *Cis*-BP v2 ^39^ using “Find Individual Motif Occurrences” (FIMO) (v5.4.1) ^61^ with default options. Next, we used CENTIPEDE (v1.2) ^40^ to call footprints for each FIMO scan result in combination with the corresponding ATAC-seq aligned bam file. Due to the presence of a large number of samples x cell types for muscle (284 samples x 13 cell types x 4,356 PWMs), we randomly picked 20 samples per cell type to perform a full scan of all available PWMs. We defined a motif occurrence to be bound by the respective TF if the CENTIPEDE posterior probability was ≥0.95 and its coordinates were fully contained within an ATAC-seq peak. We further considered any SNP overlapping such a motif occurrence to be potentially disrupting the binding site of the respective TF.

### 7. Comparison of TF ATAC-seq footprint overlapping with T2D SNPs

We combined ATAC-seq TF footprints of all motifs across samples and generated a master footprint summary file in a tissue/celltype. Next, we intersected these TF footprints with T2D 99% credible set of SNPs and identified SNPs overlapping with any TF footprints. We annotated T2D credible set of SNPs with the information of allelic imbalance and likely overlapping TF footprints. Finally, we constructed a summary counts of T2D SNPs: ATAC-seq imbalanced and overlapping with at least one TF footprint, SNPs imbalanced but not overlapping any TF footprint, SNPs not imbalanced but overlapping TF footprint and T2D SNPs not imbalanced and not footprints. We performed a hypergeometric test using R function ‘phyper’ and considered T2D imbalanced SNPs to be enriched to overlap with TF footprints for a cell type where P value < 0.05.

### 8. Enrichment of allelic imbalanced SNPs in TFBSs

We tested for enrichment of TF binding site motifs for TFs that are expressed in respective human primary tissue (average bulk TPM>0) in 15bp flanking regions around the allelic imbalanced SNPs compared to randomly shuffled input sequences as background. We used a window size of 15bp around the SNPs (including imbalanced or non-imbalanced) to assess enrichment of TFBSs accounting that the majority of TFBS motifs have size <= 15bp. We performed “Simple Enrichment Analysis” (SEA) (v5.4.1) to compare occurrences of TFBSs overlapping regions around the imbalanced SNPs compared to the background sequences ^62^. We performed this analysis for islet, liver and muscle (pseudo-bulk) as we have the most number of imbalanced SNPs (≥1,934). To account for the number of imbalanced SNPs across tissues, we selected the top 1,934 imbalanced SNPs in each tissue. We performed multiple hypothesis correction using the Benjamini-Hochberg procedure and considered motifs with FDR<0.05 to be enriched.

### 9. Enrichment of pioneer TFs overlapping imbalanced SNPs

We performed a literature search on pioneer TFs in the PubMed database (https://pubmed.ncbi.nlm.nih.gov; as of September 19, 2025) using the term “pioneer transcription factor”. We reviewed the papers and compiled a list of known pioneer TFs in 45 papers (STable 4). We tested over-representation of the pioneer TFs in the list of factors that are enriched to overlap the imbalanced SNPs in three human primary tissues in *Enrichment of allelic imbalanced SNPs in TFBSs* using hypergeometric test with R function ‘phyper’. We considered that the pioneer TFs are over-represented in the TFs inferred to be enriched to overlap the imbalanced SNPs where P value < 0.05 for each tissue.

### 10. ATAC-seq down-sampling

To assess likely tissue specificity of imbalanced SNPs, we took two layers of down-sampling approach to account for variations in number of samples with heterozygous SNPs and sequencing depth across the human primary tissues, including muscle, liver, and islet where we have the largest number of ATAC-seq imbalanced SNPs. In brief, we selected SNPs showing ASAI in any of the three tissues, and annotated them with a pool of heterozygous donors per SNP in each tissue. Next, we performed two levels of down-sampling at per SNP level. First, we selected an equal number of possible heterozygous donors with the most sequence coverage based on library size and sequencing read length across tissues while maximizing donor size (aka down-sampling at donor level). Second, we proportionally down-sampled read counts for reference and alternate allele to the tissue with the lowest coverage. We used the adjusted allelic read counts for downstream allelic imbalance using a previously described beta-binomial test. Subsequently, we performed a z-score based meta-analysis to obtain tissue-wise p values. We adjusted the P values by BH and generated a tissue-wise FDR, and considered those with FDR < 0.05 to be allelic imbalanced.

### 11. Literature review of T2D SNPs with previous functional evidence

We performed a literature review of T2D-associated SNPs with functional evidence in PubMed database (https://pubmed.ncbi.nlm.nih.gov; as of February 19, 2025) using following query.

> (“Type 2 Diabetes” OR T2D OR type 2 diabetes)

AND

> (“genetic variant” OR variant OR “SNP” OR “single nucleotide polymorphism” OR “allele-specific” OR “genetic polymorphism")

AND

> (“luciferase” OR “MPRA” OR “massively parallel reporter assay” OR “EMSA” OR “supershift” OR “ChIP-seq” OR “allelic-imbalance” OR “functional assay” OR “functional variant”)

The screening of Pubmed and EuroPMC retrieved 148 articles. We manually reviewed all papers and profiled a list of SNPs with allele-specific effect in luciferase reporter assay, EMSA, or CRISPRi/a assays. In addition, we queried SNPs with differential allelic effect with an MPRA assay ^10^ and an MPRA dataset generated from a multi-study cohort (https://mpravardb.rc.ufl.edu as of March 30, 2025). We intersected these SNPs with T2D 99% credible set of SNPs ^1^ and generated a list of T2D SNPs with previous experimental evidence.

### 12. Cell Culture

We cultured the human pancreatic β-cell line EndoC-BH1 ^63^, kindly provided by Dr. Michael Stitzel (The Jackson Laboratory), for all functional assays. We used culture flasks coated with 500X DMEM/F-12 + GlutaMAX™ (Thermo Fisher Scientific) containing 1X Fibronectin (Sigma), 5X Penicillin-Streptomycin (Thermo Fisher Scientific), and 5X Extracellular Matrix (Sigma), and incubated them for at least 1 hour at 37°C and 5% CO₂. We prepared culture medium using Advanced DMEM/F-12 (Thermo Fisher Scientific) supplemented with 50 μM 2-mercaptoethanol (Sigma), 2% BSA (Equitech-Bio), 6.7 ng/mL sodium selenite (Sigma), 5.5 μg/mL transferrin (Sigma), 2 mM GlutaMAX (Thermo Fisher Scientific) and 10 mM nicotinamide (VWR), then sterilized the medium using a 0.22 μm filter (Corning). We maintained cells at 37°C in a humidified incubator with 5% CO₂.

To passage EndoC-BH1 cells, we treated cultures with prewarmed 0.05% trypsin (Thermo Fisher Scientific) for 5 minutes at 37°C and neutralized the trypsin with an equal volume of neutralization medium consisting of 80% PBS (Thermo Fisher Scientific) and 20% heat-inactivated FBS (Bio-Techne), sterile filtered through a 0.22 μm filter (Corning). We dissociated cells by pipetting, added culture media at 3 times the trypsin volume, and counted the cells using NucleoCounter® NC-3000™ (Chemometec). We seeded cells at 80,000-90,000 cells/cm² and passaged cultures weekly.

We cultured HepG2 cells (ATCC) in DMEM (1X) + Gluta Max-1 supplemented with 4.5g/L D-glucose, 110 mg/L sodium pyruvate, 25 mM HEPES (Thermo Fisher Scientific), and 10% heat inactivated FBS (GeminiBio). To passage HepG2 cells, we treated cultures with pre-warmed 0.25% trypsin (Thermo Fisher Scientific) for 5 minutes at 37°C and neutralized the trypsin with an equal volume of culture media. We pipetted cells to dissociate, split cultures at a ratio of 1:6, and passaged cells every four days.

### 13. E*MSA*

We performed all electrophoretic mobility shift assays (EMSA) using the Odyssey® EMSA Buffer Kit (LICORBio). Nuclear extracts were obtained from EndoC-BH1 cells using NE-PER™ Nuclear and Cytoplasmic Extraction Reagents (Thermo Fisher Scientific) and protein was quantified with the Pierce™ BCA Protein Assay Kit (Thermo Fisher Scientific). Oligonucleotides comprised of 30 base pairs flanking the SNP of interest (15 upstream and 15 downstream from the SNP) were designed with IRDye700 modifications on the 5’ end (need to make a table with SNPs). Complementary forward and reverse single stranded DNA oligos for each SNP were synthesized by Integrated DNA Technologies (IDT) and annealed at 10uM concentrations.

Competitor and non-competitor unlabeled probes for the risk and non-risk alleles were added at 200X the concentration of the labeled probe for competition reactions. A pre-cast 6% native acrylamide DNA gel (Thermo Fisher Scientific) was pre-run at 100V for 30 minutes at room temperature, then placed on ice. We incubated 10X binding buffer, 1 µg/µL Poly (dl•dC), 2.5 nm labeled probes, and either 2.1 ug EndoC nuclear extract or 4 ug HepG2 nuclear extract in the dark for 20 min at room temperature, before adding 2 ul loading dye. Following incubation, 20 ul of each binding reaction was added to the gel and run at 100V on ice for 1 hour. DNA/protein binding complexes were observed through fluorescence imaging using the LICORBio Odyssey® CLx. For supershift assays, 2 ug of each antibody was added after the initial binding reaction for 15 min at room temperature.

### 14. Enhancer Cloning and Construct Generation

We selected candidate enhancer regions containing variants of interest were selected from islet- or liver-specific ATAC-seq peaks. Each enhancer sequence (∼201 bp; exact sequences listed in STable 6) was synthesized with flanking restriction enzyme cut sites by IDT using their RapidGenes service and delivered in the pUC-Ampicillin-IDT plasmid backbone. Synthesized sequences were inserted in both forward and reverse orientations. Fragments were subcloned into the pGL4.23[luc2/minP] Vector (Promega) upstream of the minimal promoter using restriction enzyme digestion and ligation. All cloned constructs were sequence-verified by PlasmidSaurus. Plasmids were transformed into NEB Stable Competent E. coli (New England Biolabs), amplified, and purified using the ZymoPURE II Plasmid Midiprep Kit (Zymo Research).

### 15. Luciferase reporter assay

We seeded EndoC-βH1 or HepG2 cells in 24-well plates at a density of 3 × 10⁵ cells per well and cultured for 48 hours before transfection. We transfected cells at 80% confluency with 1 µg of corresponding Firefly luciferase and 20 ng of Renilla luciferase control plasmid (pGL4.74[hRluc/TK], Promega) using 2 µL of Lipofectamine™ 2000 Transfection Reagent (Thermo Fisher Scientific) per well, following the manufacturer’s instructions. We measured luciferase activity at 48 hours post-transfection using the Dual-Luciferase® Reporter Assay System (Promega) on a Berthold Centro XS³ LB 960 luminometer. Firefly luciferase activity was normalized to Renilla luciferase to control for transfection efficiency, and relative transcriptional activity was reported as fold-change over empty vector control. We tested each construct in three independent biological replicates, each performed in technical triplicate. Statistical significance was assessed using a two-sided unpaired Welch’s t-test.

