## Supplementary figures and tables for "Multi-tissue analyses of allele-specific chromatin accessibility nominate likely functional variants for type 2 diabetes": Supplementary_Figure1_06-22_2025.pdf

### Supplementary figure 1

#### A Allelic imbalanced SNPs across muscle cell types

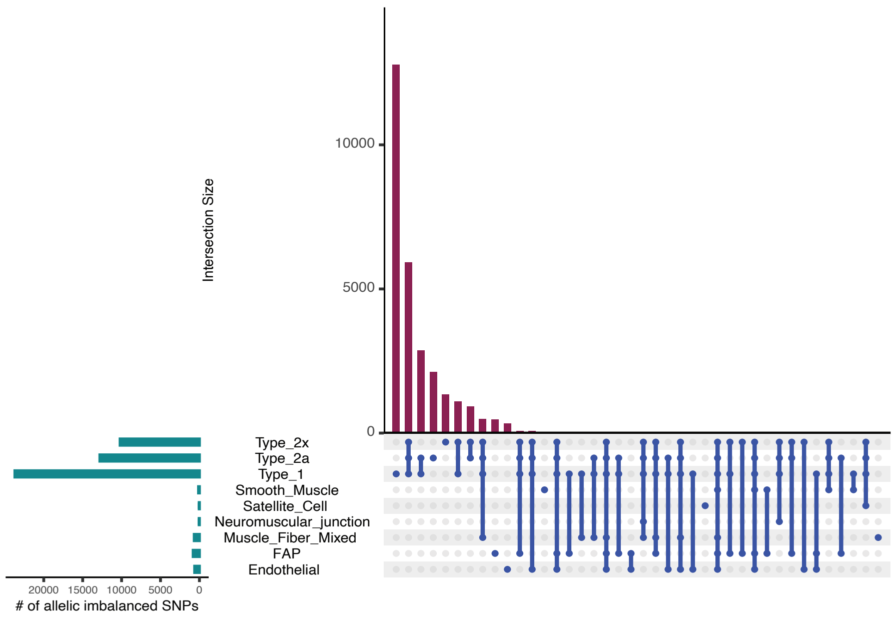


#### B Allelic imbalanced SNPs in primary human tissues before down-sampling data

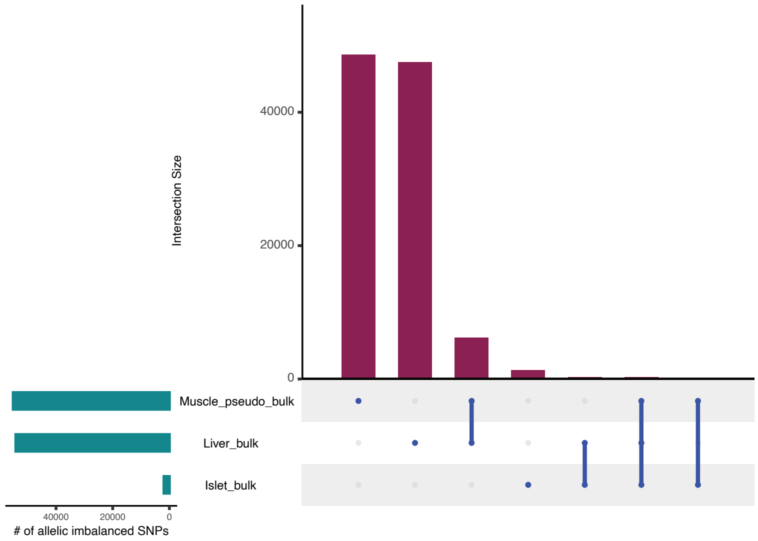
