## Supplementary figures and tables for "Multi-tissue analyses of allele-specific chromatin accessibility nominate likely functional variants for type 2 diabetes": Supplementary_Figure2_03_27_2026.pdf

### Supplementary figure 2

**A** **ASAI SNP rs13051142 was missed by the caQTL due to a moderate effect size. The same SNP is an eQTL for *LCA5L* in the same Type 1 muscle cells**

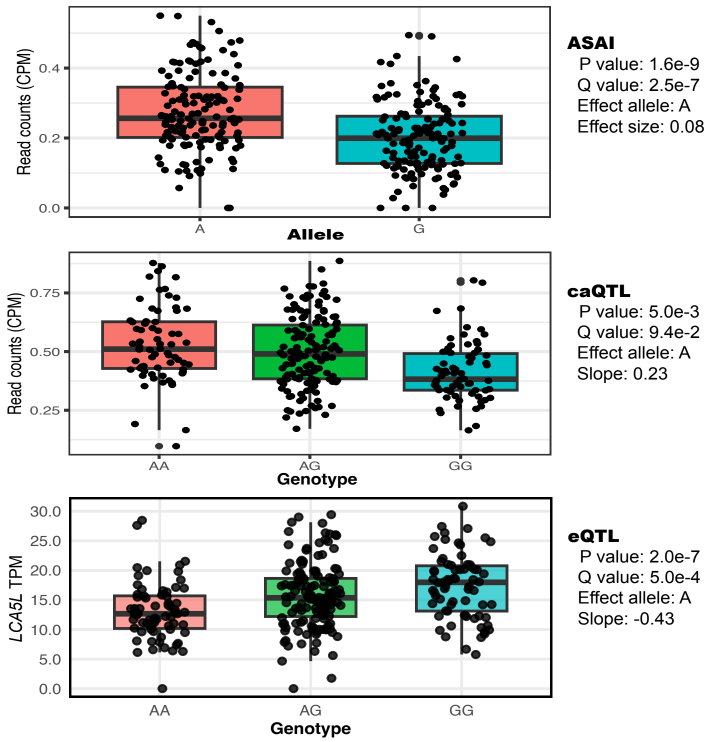


**B** **caQTl SNPs missed by ASAI tend to have lower ATAC sequencing coverage and lower MAF in Type 1 muscle cells**

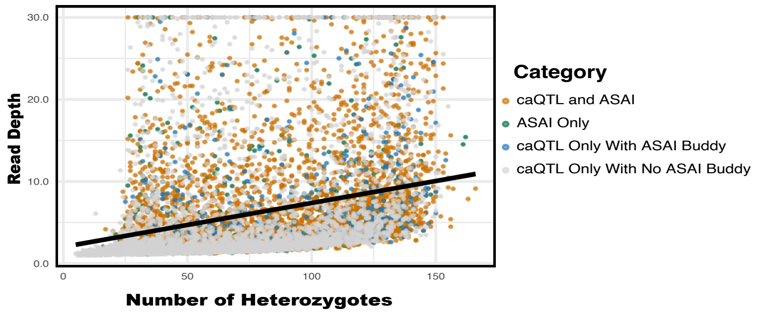


**C** **Lead caQTL SNPs missed by ASAI tend to have lower MAF in Type 1 muscle cells**

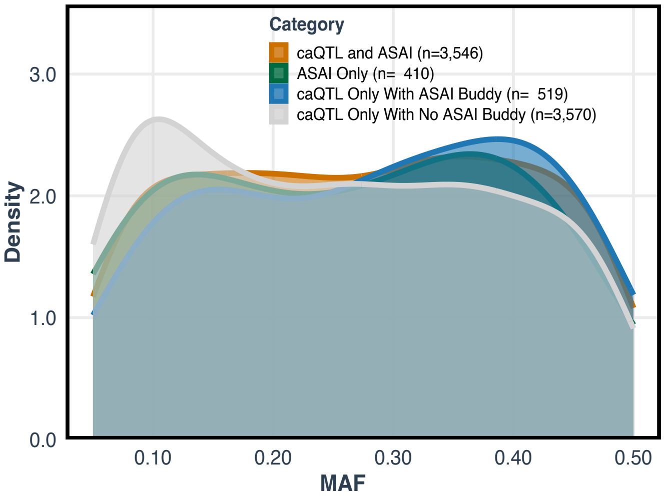
