## Supplementary figures and images for "Multi-tissue analyses of allele-specific chromatin accessibility nominate likely functional variants for type 2 diabetes"

### Supplementary_Figure3_09_08_2025.pdf

#### Supplementary figure 3

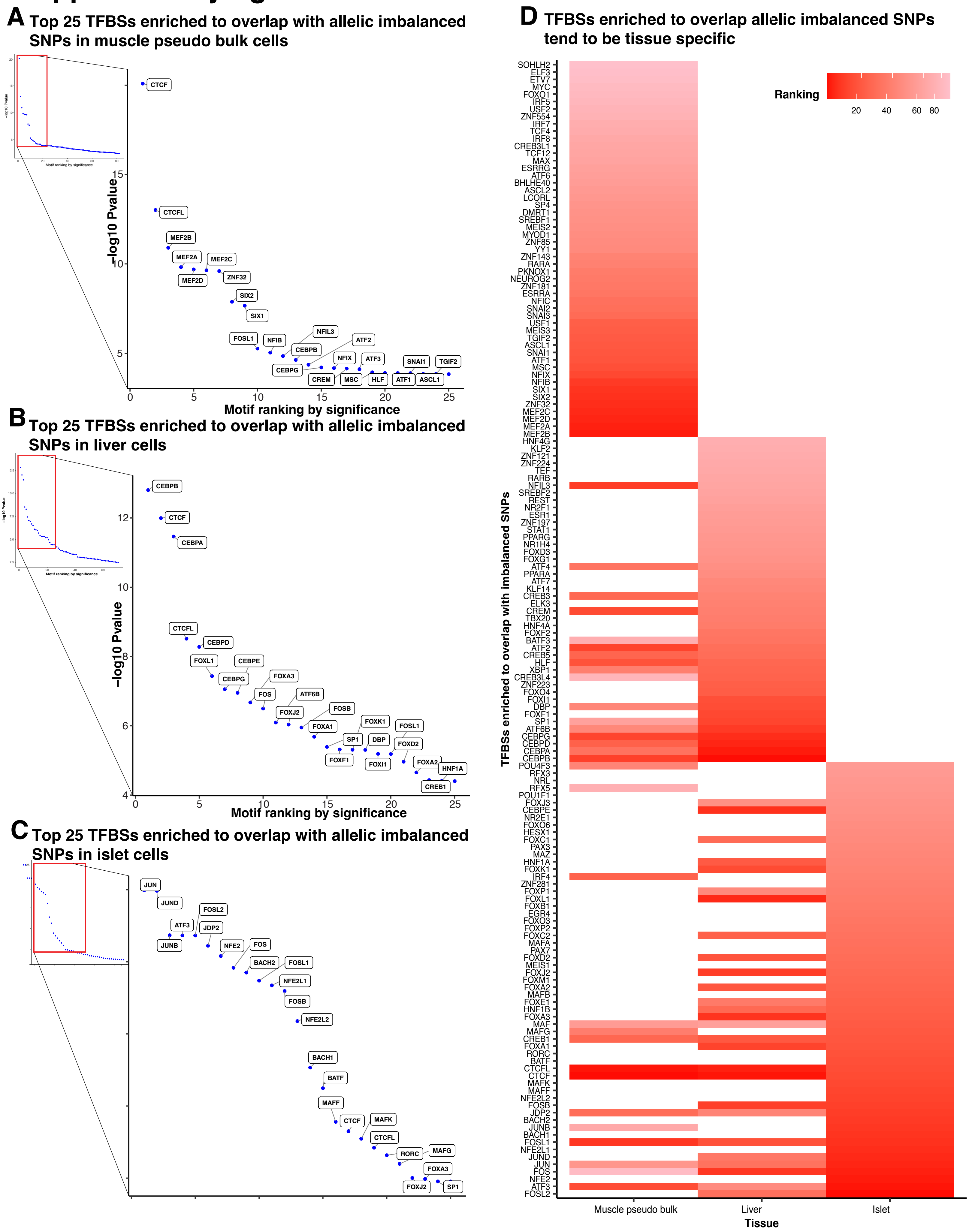

### Supplementary_Figure4_06_22_2025.pdf

#### Supplementary figure 4

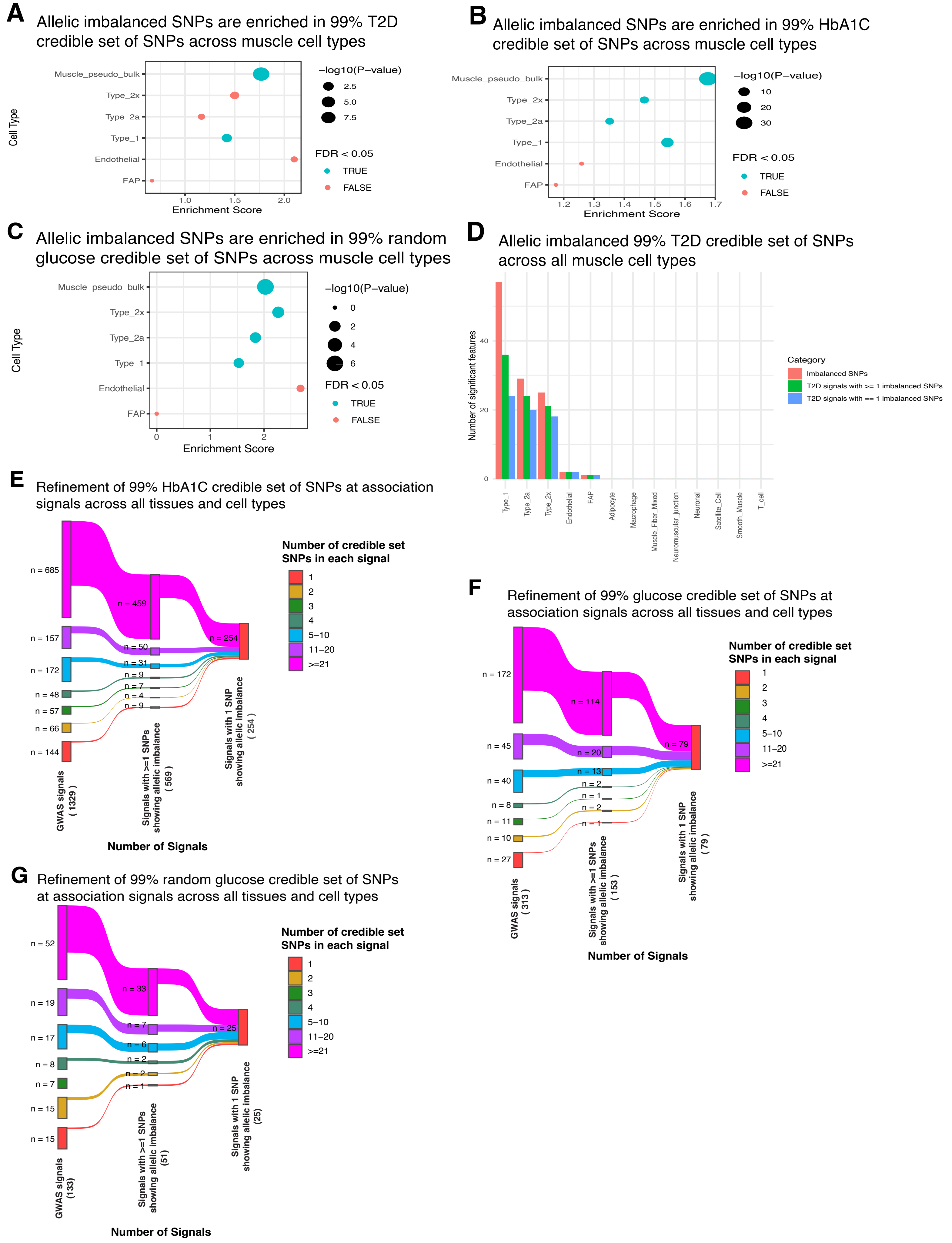

### Supplementary_Figure5_05_22_2026.pdf

#### Supplementary figure 5

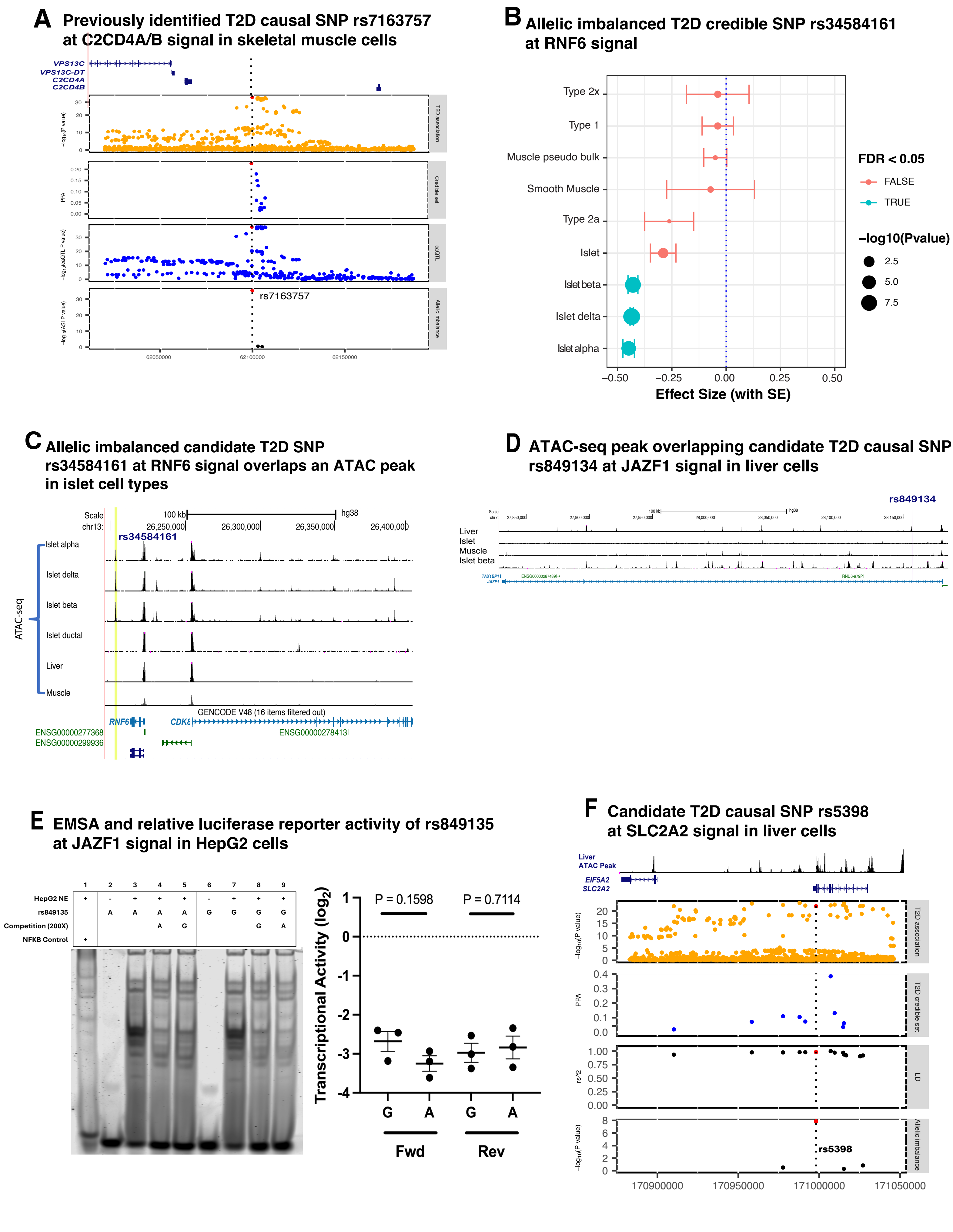
